# Evolutionary trajectories and transmission dynamics of multidrug-resistant *Mycobacterium tuberculosis* in Tibet, China

**DOI:** 10.1101/2021.02.19.21252065

**Authors:** Qi Jiang, Hai-can Liu, Qing-yun Liu, Jody E. Phelan, Li Shi, Min Gao, Xiu-qin Zhao, Jian Wang, Judith R. Glynn, Chong-guang Yang, Howard E. Takiff, Kang-lin Wan, Taane G. Clark, Qian Gao

## Abstract

**Objective:** Tibet has the highest prevalence of both tuberculosis disease and multidrug-resistant tuberculosis (MDR-TB) in China. The circulated *Mycobacterium tuberculosis* strains from Tibet were sequenced to investigate the underlying drivers for the high burden of MDR-TB.

**Methods:** Using whole-genome sequencing data of 576 *M. tuberculosis* strains isolated from consecutive patients in Tibet, we mapped resistance-conferring mutations onto phylogenetic trees to determine their evolution and spread. The impact of drug resistance on bacterial population growth was assessed with a Bayesian (Skyline Plot) analysis. Multivariable logistic regression was used to identify risk factors for the development of rifampicin resistance.

**Results:** Of the 576 isolates, 284 (49.3%), 280 (48.6%), and 236 (41.0%) were, respectively, genetically resistant to isoniazid, rifampicin, or both (MDR-TB). Among the isoniazid- and rifampicin-resistant strains, the proportions in phylogenetically-inferred clusters were 77.8% (221/284) and 62.1% (174/280), respectively. Nearly half (47.2%, 134/284) of the isoniazid-resistant strains were in six major clades, which contained between 8 and 58 strains with *katG* S315T, *katG* S315N, or *fabG1* promoter −15 C>T resistance mutations. These major clades exponentially expanded after emerging with isoniazid resistance and stabilized before evolving into MDR-TB twenty years later. Isoniazid-resistant isolates showed an increased risk of accumulating rifampicin resistance compared to isoniazid-susceptible strains, with an adjusted odds ratio of 3.81 (95% confidence interval 2.47-5.95).

**Conclusion:** Historical expansion of isoniazid-resistant strains and their increased likelihood of acquiring rifampicin resistance both contributed to the high burden of MDR-TB in Tibet, highlighting the need to detect INH-resistant strains promptly and to control their transmission.

## INTRODUCTION

Drug resistance in *Mycobacterium tuberculosis* (*M. tuberculosis*) greatly hinders the control of tuberculosis disease (TB). Approximately half a million new TB cases with resistance to rifampicin (RIF, R) occurred globally in 2019, of which 78% were also resistant to isoniazid (INH, H) and therefore multidrug-resistant (MDR)^1^. China has the second-largest percentage of global MDR-TB (14%), exceeded only by India (27%). In 2019, China had approximately 65,000 cases of MDR-TB, with high rates among both retreated (23%) and newly diagnosed TB cases (7.1%)^1^. Tibet (Xizang), an provincial autonomous region of China, has burdens of both TB and MDR-TB that are considerably higher than in other areas of China, with an estimated prevalence of TB that reached 758/100,000 population in 2014^2^. In the provincial capital city Lhasa, 21% of new and 57% of retreated cases had MDR-TB^3^.

As Tibet is relatively isolated and historically has had limited population exchange with other parts of China, it seemed feasible to trace the evolution of drug resistance and transmission dynamics that shaped its high burden of MDR-TB. To investigate the potential factors responsible for its high burden of MDR-TB, we whole-genome sequenced both susceptible and resistant Tibetan *M. tuberculosis* isolates. The resulting sequence data were used to assess the level of *M. tuberculosis* transmission, to perform a phylodynamic analysis to reconstruct the evolution of drug resistance in the local clades and to determine the impact of drug resistance on the expansion of the bacterial populations.

## METHODS

### Study site and sampling

Tibet is located on the world’s highest plateau, with an average altitude of over 4,000 metres above sea level. Its large expanse of 1.2 million square kilometres constitutes nearly an eighth of China’s entire landmass but contains only 3.3 million inhabitants (< 3 people per square-kilometre). As part of a previous study^4^, a total of 576 *M. tuberculosis* isolates were collected from consecutive patients with positive cultures at seven municipal-level clinics in Tibet over two periods ╌ during the years 2006, and 2009-2010. Genotyping of these isolates showed that the great majority (89%, 512/576) belonged to the Beijing lineage (Spoligotype ST1 strains), with limited genetic diversity^4^.

### Whole-genome sequencing and SNP calling

Genomic DNA of the *M. tuberculosis* isolates was extracted with the cetyltrimethylammonium bromide (CTAB) method^5^. A 300-base-pair double-ended DNA library of each isolate was sequenced on the Illumina HiSeq 2500 platform with an expected depth of 100-fold. Raw sequence reads were trimmed with Sickle software to remove reads of low quality (Phred base quality < 20 or read length < 10) and aligned to the H37Rv reference genome (AL123456.3) using BWA-MEM. The SAMtools software suite was applied to the alignments to call single nucleotide polymorphisms (SNPs). Fixed mutations with a frequency ≥75% were identified by VarScan. We defined genetic-clustered strains as those within a threshold genetic distance of five or fewer SNPs^6^.

### Phylogeny construction and phylodynamic analysis

The identified SNPs were used to construct a phylogeny tree using RAxML software^7^, which implemented a maximum-likelihood method with a general time-reversible model of nucleotide substitution with main node values over 0.7 from 500 bootstrapping trees. Aligned sequences were also analysed using BEAST2 software (version 2.6.1)^8^ to construct a dated phylogeny based on a molecular clock with an uncorrelated log-normal distribution and a tree prior using Coalescent Bayesian Skyline. The molecular clock was set at 1.14 (0.49-1.80) × 10^−7^ substitutions per site per year, a mutation rate reported elsewhere for lineage 2 strains^9^. Bayesian skyline plots for the effective population size were reconstructed in Tracer software^10^, with the age of the most recent common ancestor (MRCA) node being the height of the tree. The phylogenies were visualized with iTOL (version 5)^11^.

### Determination of transmitted and acquired genotypic resistance

Known drug resistance mutations with a frequency ≥ 5% in the sequence alignments were identified using the TB-profiler tool^12^ based on a published database of mutations conferring resistance to 14 anti-TB drugs^13^.

Both transmitted and acquired resistance were determined by mapping these mutations onto a phylogenetic tree^14^ and defined by the following rules: mutations shared by all strains on a branch were considered to be present at the MRCA of the branch and termed “transmitted resistance”, and these strains were considered to be phylogenetically clustered (phylo-clustered); while mutations on single branches and not present in neighbouring branches were regarded as having occurred at the terminal tip and termed “acquired resistance”. To further investigate the population growth and resistance evolution for large drug-resistant phylo-clusters, we also analysed strains on their neighbouring drug-sensitive sister branches that derived from the same ancestral nodes in the phylogeny tree.

### Data management and statistical analysis

A structured questionnaire obtained basic demographic and clinical information from enrolled patients after they signed an informed consent form, which had been reviewed and approved by the Ethical Review Board of the Chinese Center for Disease Control and Prevention (CDC). The data included: gender, age, occupation, location of residence, sampling year, BCG vaccination, TB history and smear results. The anonymized data were stored in Excel spreadsheets after double entry into EpiData (version 3.1). Univariable and multivariable logistic regression were used to assess the association of risk factors for drug resistance and to estimate the odds ratios (OR) and 95% confidence intervals (CIs). All statistical analyses were performed in R software (version 3.6.1).

## RESULTS

### Basic demographic and genetic characteristics

A total of 576 clinical isolates were collected from pulmonary TB patients in Tibet during the years 2006 and 2009-2010. More than half (53.0%, 305/576) of the samples came from TB clinics in Lhasa, the provincial capital, and the remainder from clinics in the other six administrative districts of Tibet (**<u>Suppl. Figure 1</u>**). Among 561 patients for whom the survey data were available, 325 (58.0%) were male, the median age was 32 years (interquartile range 25-43), and 288 (51.3%) reported a previous history of TB treatment (**<u>Table 1</u>**).

**Table 1.** **Basic demographic characteristics of tuberculosis patients in Tibet and the genetic characteristics of their clinical isolates, stratified by tuberculosis treatment history**

| | Total<br>N=576 | New cases<br>N=273 | Retreated cases<br>N=288 | $\chi^2$ | P value |
| --- | --- | --- | --- | --- | --- |
| Median Age (IQR), years | 32 (25-43) | 32 (23-44) | 33 (26-43) | 1.12 | 0.263 |
| Gender, n (%) |  |  |  | 4.12 | 0.042 |
| Male | 325 (58.0) | 146 (53.5) | 179 (62.2) |  |  |
| Female | 235 (42.0) | 126 (46.2) | 109 (37.8) |  |  |
| Occupation |  |  |  | 22.67 | <0.001 |
| Farmer | 298 (53.1) | 146 (53.5) | 152 (52.8) |  |  |
| Worker | 43 (7.7) | 20 (7.3) | 23 (8.0) |  |  |
| Student | 55 (9.8) | 42 (15.4) | 13 (4.5) |  |  |
| Other | 180 (31.3) | 65 (23.8) | 100 (34.7) |  |  |
| Location, n (%) |  |  |  | 9.30 | 0.002 |
| Lhasa | 305 (53.1) | 164 (60.1) | 136 (47.2) |  |  |
| Other | 271 (47.0) | 109 (39.9) | 152 (52.8) |  |  |
| Sample year, n (%) |  |  |  | 0.46 | 0.794 |
| 2006 | 177 (30.7) | 81 (30.0) | 82 (28.5) |  |  |
| 2009 | 172 (30.0) | 80 (29.3) | 92 (31.9) |  |  |
| 2010 | 227 (39.4) | 112 (41.0) | 114 (39.6) |  |  |
| Smear status |  |  |  | 12.29 | 0.006 |
| Negative or scanty | 245 (42.5) | 137 (50.2) | 108 (37.5) |  |  |
| 1+ | 142 (25.3) | 68 (24.9) | 74 (25.7) |  |  |
| 2+ | 94 (16.8) | 34 (12.5) | 60 (20.8) |  |  |
| 3+ | 80 (14.3) | 34 (12.5) | 46 (16.0) |  |  |
| Lineage, n (%) |  |  |  | 0.26 | 0.878 |
| Lineage 2 | 524 (91.0) | 247 (90.5) | 264 (91.7) |  |  |
| Lineage 4 | 36 (6.3) | 18 (6.6) | 17 (5.9) |  |  |
| Lineage 3 | 16 (2.8) | 8 (2.9) | 7 (2.4) |  |  |
| Genetic resistance, n (%) |  |  |  | 106.99 | <0.001 |
| Pan-susc. | 231 (40.1) | 160 (58.6) | 67 (23.3) |  |  |
| DR | 109 (18.9) | 61 (22.3) | 47 (16.3) |  |  |
| MDR | 187 (32.5) | 42 (15.4) | 136 (47.2) |  |  |
| Pre-XDR | 41 (7.1) | 10 (3.7) | 31 (10.8) |  |  |
| XDR | 8 (1.4) | 0 (0) | 7 (2.4) |  |  |
| Genetic clustering, n (%) |  |  |  |  |  |
| 1-SNP | 105 (18.2) | 44 (16.1) | 57 (19.8) | 1.28 | 0.258 |
| 5-SNP | 216 (37.5) | 93 (34.1) | 118 (41.0) | 2.85 | 0.091 |
| 12-SNP | 339 (58.9) | 153 (56.0) | 176 (61.1) | 1.48 | 0.223 |
| 25-SNP | 444 (77.1) | 208 (76.2) | 222 (77.1) | 0.06 | 0.803 |
| 50-SNP | 509 (88.4) | 238 (87.2) | 257 (89.2) | 0.57 | 0.450 |

Abbreviations: IQR=interquartile range; Pan-susc. = pan-susceptible; DR = Drug Resistant but not MDR-TB; MDR-TB = MultiDrug-Resistant tuberculosis; XDR = eXtensively Drug-Resistant; SNP = single nucleotide polymorphism.
Note: Demographic data (tuberculosis treatment history, age and gender) was missing for 16 cases.

The whole-genome sequences of the 576 *M. tuberculosis* clinical isolates had an average depth of 79-fold (range 22-to 137-fold). A total of 2,433,862 SNPs were identified, of which 1,389 were resistance-conferring mutations across 14 anti-TB drugs. A maximum-likelihood phylogeny tree constructed using all SNPs (**<u>Figure 1</u>**) revealed that the great majority of isolates (524, 91.0%) belonged to Beijing Lineage 2, with a minority belonging to the Euro-American Lineage 4 (36, 6.3%) or Central Asian Lineage 3 (16, 2.8%). From the genotypic resistance profiles, 231 (40.1%) isolates were pan-susceptible while the other 345 (59.9%) strains were resistant to at least one anti-TB drug, including 236 (41.0%) that were MDR-TB. The proportions of new and retreated patients with MDR-TB were 19.0% (52/273) and 60.4% (174/288), respectively (**<u>Table 1)</u>**.

**Figure 1.**
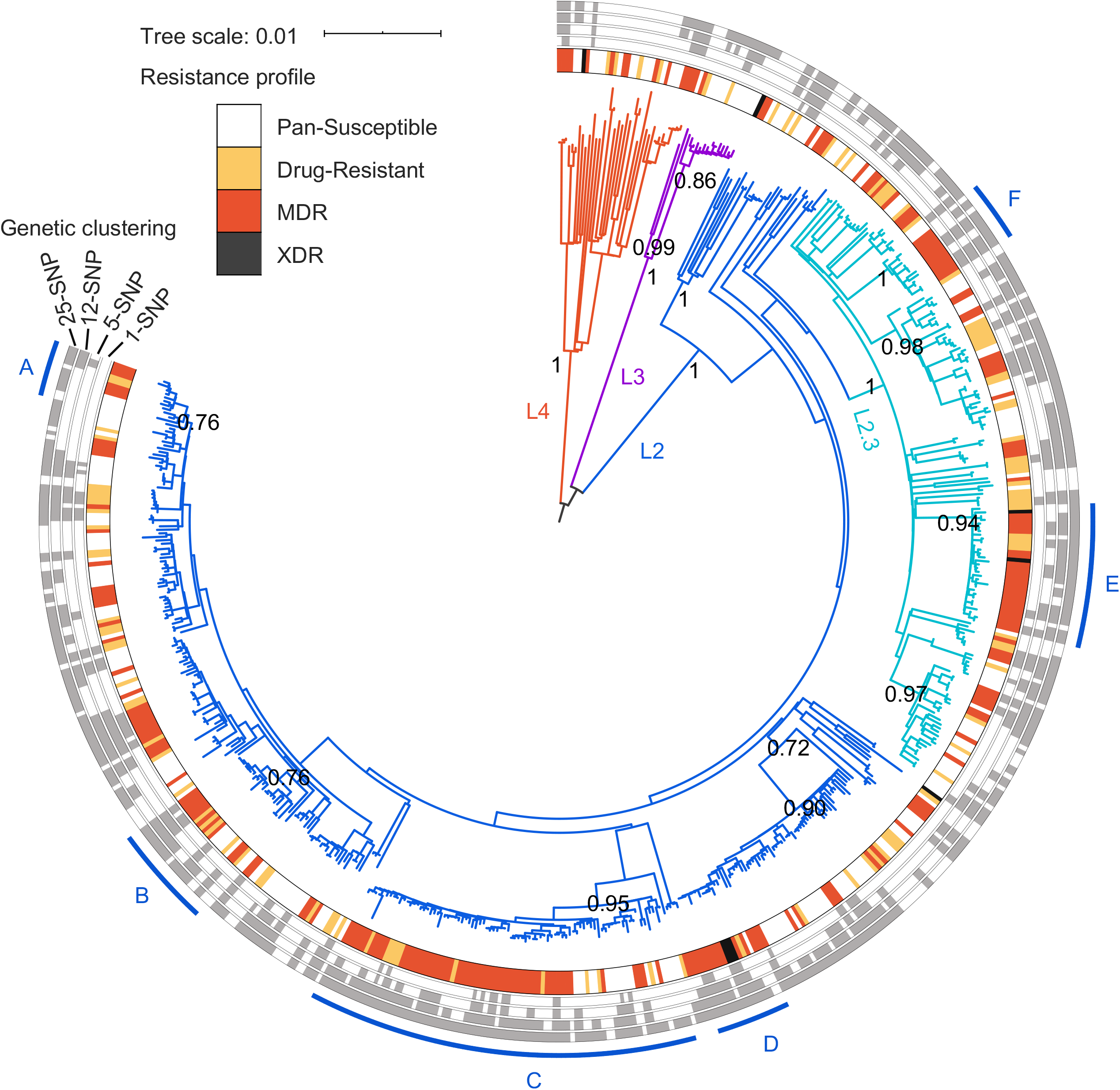
Genetic structure of prevalent *M. tuberculosis* in Tibet. The maximum-likelihood phylogeny tree of 576 *M. tuberculosis* clinical isolates sampled during years 2006 and 2009-2010 in Tibet, was rooted at midpoint with key nodes gaining a bootstrap value over 0.7. Bootstrap values are shown as numbers on major nodes. Coloured branches show three identified lineages: Beijing strains (Lineage 2, L2) in blue, (Modern Beijing sub-lineage L2.3 in light blue), Central Asian Strain (CAS) (L3) in purple, and Euro-American lineage (L4) in orange. The innermost surrounding circle indicates the genotypic resistance profile of each strain, including pan-susceptible (white), drug-resistant (yellow), MDR (orange), and XDR strains (black). The outermost surrounding circles indicate genetic clustering with thresholds of 1-, 5-, 12-, and 25-SNP distances. Major drug-resistant clades are labelled A-F.

Although many studies have used thresholds between 5 and 12 SNPs to define a genetic cluster^6,15^, the Tibet strains showed no clear separation but rather a gradual decrease in the distribution of the SNP differences between strains (**<u>Figure 2</u>**). Using a limit of 5 SNPs to define clustered strains, Lineage 2 strains were nearly twice as likely to be in genetic clusters (39.3%, 206/524) as the few Lineage 4 strains collected (22.2%, 8/36); and MDR-TB strains showed significantly more clustering (47.0%, 111/236) than either pan-susceptible strains (31.2%, 72/231) or those with other resistance profiles (30.3%, 33/109) (*P*<0.001) (**<u>Figure 2</u>**). The resistance profiles among different lineages did not show a significant difference (**<u>Suppl. Table 1</u>**).

**Figure 2.**
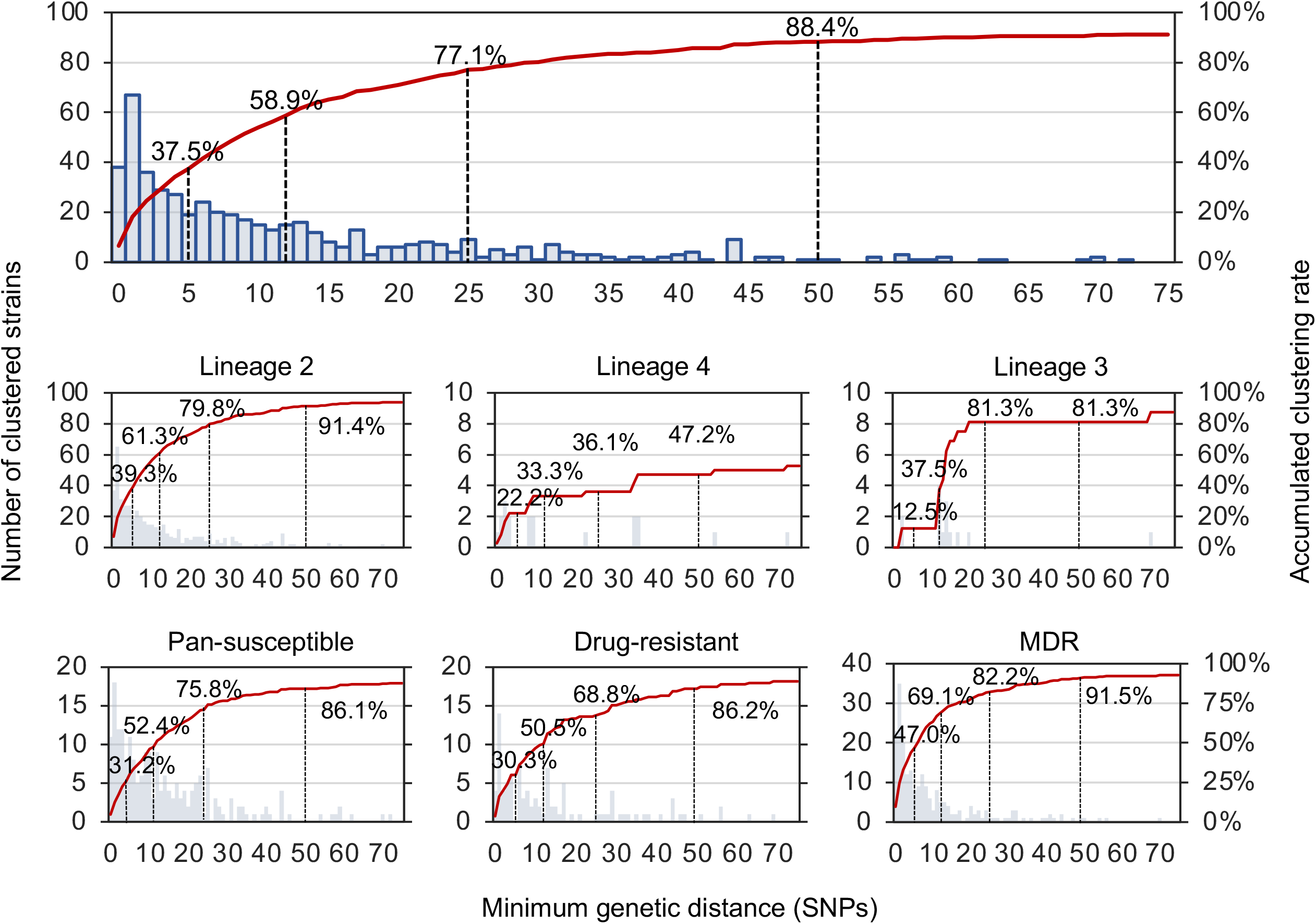
Distribution of minimum genetic distance and accumulated clustering rates overall, and stratified by strain lineage and drug resistance profile. Vertical lines indicate the proportions of strains in 5-SNP, 12-SNP, 25-SNP and 50-SNP clusters, respectively. “Drug-resistant” means drug-resistant tuberculosis but not MDR-TB. Most strains (88.4%, 509/576) were separated from another strain by no more than 50 SNPs, and the proportions of strains separated by up to 5, 12 or 25 SNPs from another isolate were 27.5%, 58.9% and 77.1%, respectively.

### Transmitted drug resistance

The mutations conferring drug resistance in *M. tuberculosis* strains have different fitness costs that can affect their transmissibility^16^. Most INH-resistant strains carried the low-fitness cost *katG* S315T mutation and showed a clustering rate of 41.8% (74/177), using the 5-SNP limit. Although strains carrying other INH-resistance mutations were much less common, they all had higher clustering rates than the pan-sensitive strains, except for those with the *katG* S315N mutation (**<u>Table 2</u>**). Among 280 RIF-resistant strains, the most frequent mutation was the low-fitness cost *rpoB* S450L (99, 35.4%). The next most common was the *rpoB* H445Y (42, 15.0%) mutation and there were a few strains with two other substitutions of the same amino acid, all of which had higher clustering rates (*rpoB* H445Y/D/R, combined 51.3% [41/80]) than strains with *rpoB* S450L (37.4% [37/99]) (**<u>Table 2</u>**).

**Table 2.**
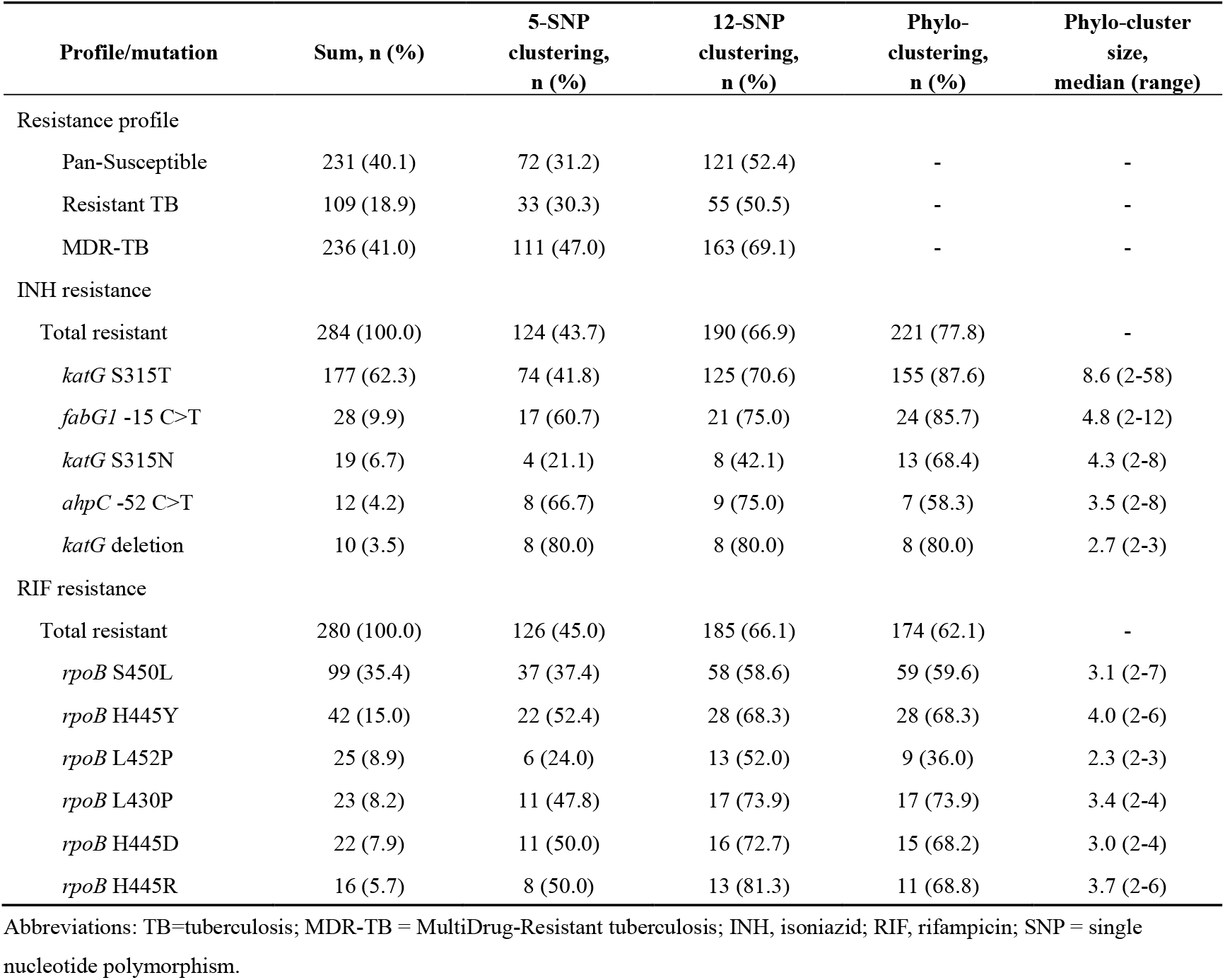
**Genetic and phylogenetic clustering of *M. tuberculosis* strains with the principal mutations conferring isoniazid (INH) or rifampicin (RIF) resistance**

| Profile/mutation | Sum, n (%) | 5-SNP<br>clustering,<br>n (%) | 12-SNP<br>clustering,<br>n (%) | Phylo-<br>clustering,<br>n (%) | Phylo-cluster<br>size,<br>median (range) |
| --- | --- | --- | --- | --- | --- |
| Resistance profile |  |  |  |  |  |
| Pan-Susceptible | 231 (40.1) | 72 (31.2) | 121 (52.4) | - | - |
| Resistant TB | 109 (18.9) | 33 (30.3) | 55 (50.5) | - | - |
| MDR-TB | 236 (41.0) | 111 (47.0) | 163 (69.1) | - | - |
| INH resistance |  |  |  |  |  |
| Total resistant | 284 (100.0) | 124 (43.7) | 190 (66.9) | 221 (77.8) | - |
| *katG* S315T | 177 (62.3) | 74 (41.8) | 125 (70.6) | 155 (87.6) | 8.6 (2-58) |
| *fabG1* -15 C>T | 28 (9.9) | 17 (60.7) | 21 (75.0) | 24 (85.7) | 4.8 (2-12) |
| *katG* S315N | 19 (6.7) | 4 (21.1) | 8 (42.1) | 13 (68.4) | 4.3 (2-8) |
| *ahpC* -52 C>T | 12 (4.2) | 8 (66.7) | 9 (75.0) | 7 (58.3) | 3.5 (2-8) |
| *katG* deletion | 10 (3.5) | 8 (80.0) | 8 (80.0) | 8 (80.0) | 2.7 (2-3) |
| RIF resistance |  |  |  |  |  |
| Total resistant | 280 (100.0) | 126 (45.0) | 185 (66.1) | 174 (62.1) | - |
| *rpoB* S450L | 99 (35.4) | 37 (37.4) | 58 (58.6) | 59 (59.6) | 3.1 (2-7) |
| *rpoB* H445Y | 42 (15.0) | 22 (52.4) | 28 (68.3) | 28 (68.3) | 4.0 (2-6) |
| *rpoB* L452P | 25 (8.9) | 6 (24.0) | 13 (52.0) | 9 (36.0) | 2.3 (2-3) |
| *rpoB* L430P | 23 (8.2) | 11 (47.8) | 17 (73.9) | 17 (73.9) | 3.4 (2-4) |
| *rpoB* H445D | 22 (7.9) | 11 (50.0) | 16 (72.7) | 15 (68.2) | 3.0 (2-4) |
| *rpoB* H445R | 16 (5.7) | 8 (50.0) | 13 (81.3) | 11 (68.8) | 3.7 (2-6) |

Abbreviations: TB=tuberculosis; MDR-TB = MultiDrug-Resistant tuberculosis; INH, isoniazid; RIF, rifampicin; SNP = single nucleotide polymorphism.

Mapping the resistance-conferring mutations on the phylogenetic tree showed that 77.8% (221/284) of strains resistant to INH and 62.1% (174/280) resistant to RIF, belonged to phylo-clusters that resulted from the transmission of resistance mutations from parental strains (**<u>Table 2</u>**). Even higher proportions of strains resistant to SM (82.7%, 191/231) or PAS (96.0%, 72/75) resulted from transmitted resistance, as well as over half of strains resistant to the other first-line drugs ethambutol (EMB, E) (57.1%, 97/170) and pyrazinamide (PZA, Z) (52.1%, 63/121) (**<u>Suppl. Table 2</u>**). The sizes of the INH-resistant phylo-clusters were generally larger than those with RIF resistance (**<u>Table 2</u>**). There were four large phylo-clusters of strains with the *katG* S315T mutation, each containing more than ten strains, while the RIF resistant clusters had, at most, seven strains. To reconstruct the growth history of the bacterial population and the trajectories of resistance accumulation, we focused on six large phylo-clusters that contained more than eight strains. In total, these six large phylo-clusters included nearly half of the INH-resistant isolates (47.2%, 134/284) or of the MDR-TB strains (48.7%, 115/236). For further analysis, each of these six phylo-clusters, together with their sister branches, were considered as clades, labelled A-F (**<u>Figure 1)</u>**.

### Evolutionary trajectories of drug resistance in the major clades

To determine when these six clades developed antibiotic resistance to each drug, we constructed dated phylogenetic trees for each, and estimated the evolutionary time to the appearance of nodes whose offspring strains all harboured the same resistance-conferring mutations. All of these clades first evolved INH resistance conferred by acquiring *katG* S315T (Clades B-E), *katG* S315N (Clade A) or *fabG1* promoter −15 C>T (Clade F) mutations, and subsequently evolved into phylo-clusters containing between 8 and 58 INH-resistant strains (**<u>Table 3</u>**). In five of these six clades, the mutations conferring INH resistance were acquired around year 1970 (95% CI 1967-1975), and the remaining Clade D acquired the *katG* S315T around 1989 (95% CI 1980-1998). Three clades (B, D and E) also acquired SM resistance at around the same time as INH resistance, and one clade (C) acquired additional resistance to PAS (**<u>Figure 3; Suppl. Figure 2</u>**).

**Table 3.** Accumulation of mutations conferring drug resistance for anti-tuberculosis drugs among the six major clades A-F.

| Clade | Initial resistance mutation | Estimated year of first INH resistance<br>(95% credible intervals) | # phylo-clustered strains (+ # sister branches) | # strains with resistance / # mutation accumulation events |  |  |  |  |  |  |  | Resistant to # drugs per strain |
| --- | --- | --- | --- | --- | --- | --- | --- | --- | --- | --- | --- | --- |
|  |  |  |  | INH | SM | PAS | RIF | EMB | PZA | FQ | SLD |  |
| Clade A | *katG* S315N | 1972.81 (1957.70-1986.26) | 8 (+4) | 8/1 | 7/4 | 0 | 6/4 | 3/2 | 2/2 | 0 | 0 | 3.3 |
| Clade B | *katG* S315T | 1975.36 (1961.07-1987.82) | 12 (+10) | 12/2 | 12/1 | 2/1 | 9/4 | 9/3 | 6/1 | 3/2 | 0 | 4.8 |
| Clade C | *katG* S315T | 1967.55 (1957.58-1977.10) | 58 (+26) | 58/1 | 58/2 | 58/1 | 51/23 | 35/13 | 21/11 | 8/6 | 0 | 5.0 |
| Clade D | *katG* S315T | 1989.69 (1980.47-1998.02) | 13 (+3) | 13/1 | 13/1 | 4/2 | 11/4 | 3/4 | 4/1 | 4/1 | 3/1 | 4.2 |
| Clade E | *katG* S315T | 1973.72 (1964.21-1983.33) | 31 (+0) | 31/1 | 31/1 | 0 | 26/14 | 16/11 | 7/5 | 7/6 | 4/3 | 4.2 |
| Clade F | *fabG1* promoter -15 C>T | 1969.29 (1954.49-1982.62) | 12 (+1) | 12/4 | 3/3 | 0 | 12/9 | 10/9 | 6/3 | 0 | 3/3 | 4.2 |

Abbreviations: INH, isoniazid; SM, streptomycin; PAS, P-aminosalicylic acid; RIF, rifampicin; EMB, ethambutol; PZA, pyrazinamide; FQ, fluoroquinolones; SLD, second-line injectable drugs (amikacin, kanamycin, capreomycin).

**Figure 3.**
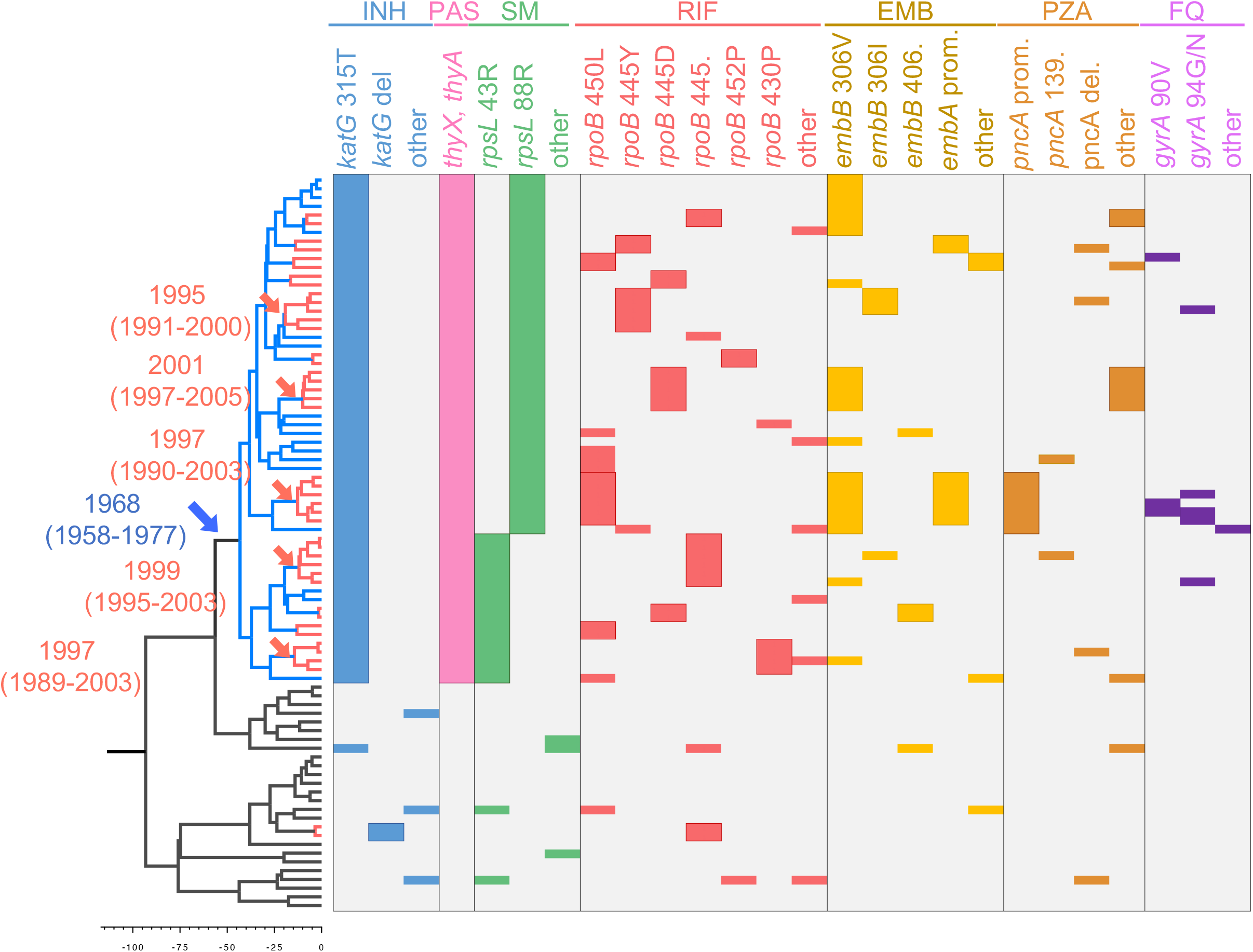
Determination of acquisition or transmission of resistance-conferring mutations. The left side shows a dated phylogeny tree of Clade C, with blue and red branches indicating transmitted resistance to INH and RIF, respectively. The right side shows various resistance mutations in different panels with colors indicating resistance to the corresponding drugs, as labelled above. The blue arrow points to nodes where INH resistance mutations occurred with the approximate dates in blue, while red arrows and dates indicate the occurrence of RIF-resistance mutations transmitted to three or more strains. Abbreviations: INH, isoniazid; SM, streptomycin; PAS, P-aminosalicylic acid; RIF, rifampicin; EMB, ethambutol; PZA, pyrazinamide; FQ, fluoroquinolones

In all six clades, RIF resistance developed after 1990, much later than INH resistance and consistent with the time period in which RIF became widely used for treating TB in Tibet. Notably, each clade contained strains with different RIF-resistance-conferring mutations, with no single mutation being dominant. Similarly, nearly all clades contained more than one mutation conferring resistance to the other first-line drugs EMB and PZA. After these clades developed INH resistance, mutations conferring RIF resistance occurred between 4 and 23 times independently in different sub-clades, while EMB and PZA resistance occurred 2 to 13 times and 1 to 11 times, respectively (**<u>Table 3</u>**). Amongst all the sampled isolates, we identified 110 independent mutational events for INH resistance and 161 for RIF resistance, among which 31.8% (35/110) and 34.8% (56/161), respectively, were transmitted to secondary strains in the phylo-clusters (**<u>Suppl. Table 2</u>**)

Taking the largest clade, Clade C, as an example to illustrate the evolutionary trajectories in detail (**<u>Figure 3</u>**), all 58 phylo-clustered strains of Clade C carried the same mutations for INH (*katG* S315T) and PAS resistance (*thyX* promoter −16 C>T and *thyA* large segment deletion), both of which were estimated to have occurred around year 1968 (95% CI 1958-1977). Resistance to SM was subsequently acquired around year 1972, with simultaneous mutations of either *rpsL* K88R (1963-1980) or *rpsL* K43R (1962-1981), creating two sub-clades resistant to all drugs in the triple TB therapy used at the time (INH, SM and PAS). The mutations conferring resistance to RIF occurred more than 20 years later on at least 23 descendent branches of this clade, resulting in 12 RIF-resistant phylo-clusters of 2-6 strains, and another 11 non-clustered RIF-resistant strains (**<u>Figure 3</u>**), indicating both acquired and transmitted resistance. Mutations conferring resistance to EMB and PZA also developed independently on multiple branches, producing 19 strains that were resistant to all four of the first-line drugs used in the standard HRZE therapy. Few strains belonging to this clade developed resistance to second-line anti-TB drugs, including only eight pre-XDR-TB strains with fluoroquinolones-resistant mutations in *gyrA* (90V, 94G, 94N) or *gyrB* (461N). The other clades underwent similar trajectories, acquiring mutations conferring resistance to RIF and other first-line drugs multiple times in parallel in the descendants of strains with pre-existing INH resistance (**<u>Suppl. Figure 2</u>**).

### Risk of INH resistance for population expansion and resistance accumulation

To evaluate the impact of INH and RIF resistance on the clonal expansion of local TB strains, we used Bayesian skyline plots to reconstruct the growth of the bacterial populations in the six major clades (**<u>Figure 4</u>**). Before the emergence of drug resistance, mainly resistance to INH, the bacterial population of these clades showed varied patterns of expansion or shrinkage. Irrespective of their previous patterns however, after the development of INH resistance there followed a period of exponential growth during which the bacterial populations of these clades expanded for decades. Subsequently, after the acquisition of RIF resistance, the population of most clades stabilized (Clade E) or decreased (Clades A, B, D, F) (**<u>Figure 4</u>**), and only Clade C continued to expand while acquiring many RIF-resistance mutations independently on multiple branches.

**Figure 4.**
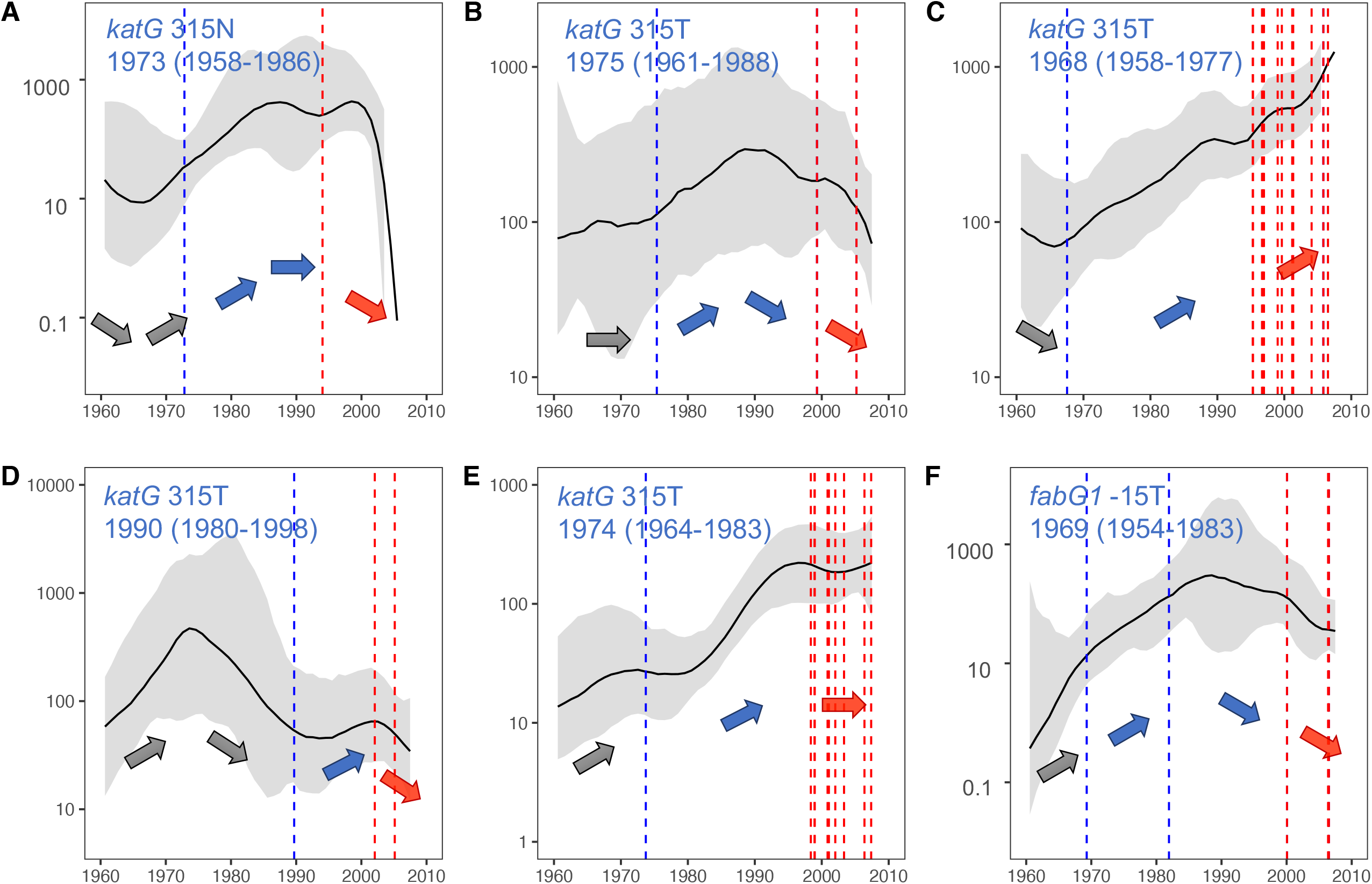
The effective population size of the six large clades A-F. Blue and red dash lines indicating the acquisition of isoniazid (INH) and rifampicin (RIF) mutations, respectively. Note, the scales vary.

We suspected that strains with pre-existing INH resistance would be more likely to accumulate mutations conferring RIF resistance than INH-sensitive strains. For example, 87.9% (51/58) of the INH-resistant Clade C strains acquired RIF resistance, compared to only 19.2% (5/26) of strains on its sister INH-sensitive branches (**<u>Figure 3</u>**). To quantify the effect of pre-existing INH resistance on the accumulation of RIF resistance, we tested it as an independent variable in a logistic regression model with the whole dataset. In the univariable analysis, the development of RIF resistance was significantly associated with strains that had pre-existing INH resistance, belonged to a genetic cluster (genetic distance ≤ 5 SNPs), were isolated from retreated patients, were isolated in Lhasa, or were sampled in 2006 rather than in 2009/2010 (**<u>Table 4</u>**). These variables were then included in a multivariable logistic regression, adjusting for age and gender and excluding cases with missing data. The final model showed that, *M. tuberculosis* strains with pre-existing INH resistance had an adjusted odds ratio of developing RIF resistance of 3.81 (95% CI: 2.47-5.95) compared to INH-susceptible strains, and genetic-clustered strains within a 5-SNP distance had an adjusted odds ratio of 2.08 (95% CI:1.37-3.18) compared to non-clustered strains (**<u>Table 4</u>**).

**Table 4.** Risk factors for the development of rifampicin (RIF) resistance in univariable and multivariable logistic regression models.

|  | RIF-resistant strains<br>N=280, m±sd/n (%) | RIF-sensitive strains<br>N=296, m±sd/n (%) | Univariable regression |  | Multivariable regression |  |
| --- | --- | --- | --- | --- | --- | --- |
|  |  |  | Unadjusted odds ratio (95%CI) | P value | Adjusted odds ratio (95%CI) | P value |
| Age, years* | 33.8 ± 14.7 | 34.7 ±15.6 | 1.00 (0.99-1.01) | 0.481 | 1.00 (0.99-1.01) | 0.899 |
| Male Gender* | 160 (59.5) | 165 (56.7) | 1.12 (0.80-1.57) | 0.506 | 0.93 (0.61-1.41) | 0.731 |
| Sampled in Lhasa | 127 (45.4) | 173 (58.4) | 0.62 (0.44-0.86) | 0.004 | 0.72 (0.48-1.08) | 0.109 |
| Sampled in 2006 | 98 (35.0%) | 79 (26.7) | 1.48 (1.04-2.11) | 0.031 | 1.74 (1.11-2.74) | 0.016 |
| Clustering ≤ 5 SNPs | 126 (45.0) | 90 (30.4) | 1.87 (1.33-2.64) | <0.001 | 2.08 (1.37-3.18) | 0.001 |
| Pre-existing isoniazid resistance | 135 (48.2) | 58 (19.6) | 3.82 (2.65-5.56) | <0.001 | 3.81 (2.47-5.95) | <0.001 |
| TB history* | 205 (76.2) | 83 (28.4) | 8.07 (5.55-11.85) | <0.001 | 8.14 (5.43-12.38) | <0.001 |

Note: \*Demographic data was missing for 16 cases, which were excluded from the multivariable regression.

## DISCUSSION

In this study of *M. tuberculosis* isolates from Tibet, China, we characterized evolutionary trajectories for MDR-TB and demonstrated a high level of transmitted resistance to both INH and RIF drugs. However, six clades were shown to expand exponentially after acquiring INH-resistance mutations in the 1970s, and mostly stabilized before accumulating multiple RIF-resistance mutations to become MDR-TB in the late 1990s. Compared to INH-susceptible strains, strains with mutations conferring INH resistance were found to have a significantly higher risk of acquiring RIF resistance. Both the historical expansion of INH-resistant strains and the increased risk of these strains acquiring additional RIF resistance likely contributed to the current high burden of MDR-TB in Tibet.

The rate of genotypic drug-resistance among sampled *M. tuberculosis* isolates in Tibet was extremely high, with 59.9% of isolates resistant to any anti-TB drug, and 41.0% MDR-TB. Recent transmission contributed almost half (47.0%) of the MDR-TB cases, using the genetic-clustering threshold of 5 SNPs. This proportion increased to 69.1% when the genetic-clustering threshold was set at 12 SNPs. The level of recent transmission of MDR-TB in Tibet was much higher than that in other areas of China, such as Shanghai^15^ or Shenzhen^17^ (32% and 25%, respectively, with a 12-SNP threshold,), and was similar to the proportions of MDR-TB attributed to recent transmission in other countries with high MDR-TB burdens, such as Russia (60% with terminal branch length of ≤ 5 SNPs)^14^ or South Africa (79% in phylogenetic clades)^18^. However, while the majority of the MDR-TB burden in these other countries is caused by a few MDR strains that are extensively transmitted, most MDR-TB strains in Tibet belonged to small genetic-clusters of only 2 to 6 cases. We thus speculate that the high prevalence of MDR-TB in Tibet is more likely the result of poor TB control than the high transmissibility of particular strains. Due to relatively poor healthcare infrastructure in Tibet, the proportion of TB cases tested for drug resistance was low. The Tibet CDC reported that between July 2017 and June 2018, only 8 out of 476 (1.8%) smear-positive TB cases were tested for drug sensitivity and just one MDR-TB case was diagnosed. The underdiagnosis of MDR-TB in Tibet could result in inappropriate treatment regimens, poor treatment outcomes and prolonged transmission of MDR-TB strains.

We could phylogenetically follow the progression from INH-resistant TB to MDR-TB through the acquisition of mutations conferring RIF resistance, especially within the six major clades. We showed that strains with pre-existing INH resistance were more likely to develop RIF resistance, which is consistent with an *in vitro* experimental study^19^ that showed that laboratory-generated INH-resistant strains were more likely to accumulate RIF-resistant mutations than INH-sensitive strains. In a real-world dataset, a recent meta-analysis similarly found that treatment of INH-resistant, RIF-sensitive strains with the standard first-line HRZE regimen resulted in high proportions of unfavourable outcomes, including relapse or treatment failure rate (15%) and development of MDR-TB (3.6%)^20^. A recent study^21^ highlighted the high global prevalence of INH-resistant TB, with proportions among new and retreated patients of 7.4% and 11.4%, respectively. If undetected and treated with ineffective regimens, these numerous INH-resistant strains could be transmitted during prolonged infections, with an increased likelihood of acquiring further resistance.

INH resistance is thought to evolve earlier than resistance to the other first-line drugs. A study^22^ using a large global dataset of *M. tuberculosis* whole-genome sequences found that the INH-resistance mutation *katG* S315T overwhelmingly arose before RIF resistance across different lineages and regions. In our study, four of the six large clades were found to have first acquired the *katG* S315T mutation, and one clade each acquired the *katG* S315N and *fabG1* promoter −15 C>T mutations. The INH resistance impaired the effectiveness of triple therapy that was being prescribed at the time, and from the 1970s to 1990s these clades evolved into large phylo-clusters before RIF, EMB and PZA were introduced into TB treatment in Tibet. From the phylogenetic trees, we could see that the mutations conferring RIF resistance then occurred independently, in parallel, on multiple branches of these INH-resistant clades, and subsequently disseminated to cause large numbers of MDR-TB cases. This pattern differs from several MDR-TB outbreaks reported elsewhere that were caused by the extensive transmission of a very few dominant MDR-TB strains^14,23^. In a huge outbreak over four decades in Argentina^24^, RIF resistance was acquired soon after the development of INH resistance. In London^25^, an MDR-TB outbreak was found to have originated from an INH-resistant strain, but RIF resistance was acquired only once during transmission. The specific pattern for multiple, independent acquisitions of RIF resistance among *M. tuberculosis* in Tibet may be a result of the historical expansion of INH-resistant strains before the implementation of RIF.

Half of the samples included in this study came from retreated TB patients, a proportion higher than the usual rate, but the overall retreatment rate of TB in Tibet is much higher than in other regions of China. According to a cross-sectional investigation by Tibet CDC in 2014^26^, the proportion of retreated patients reached 37.5% in Lhasa, and averaged 26.6% in other Tibetan cities. The high percentage of retreated patients in this study could also partially result from sampling bias at the municipal general hospitals, where the cultures available for this study were more likely to have been obtained from severe and refractory cases. Correspondingly, the drug-resistance rates in this study are also higher than the average resistance rates in Tibet. However, the overrepresentation of resistant strains should not affect our characterization of the evolution of drug resistance, the history of bacterial population expansion and the contribution of drug resistant strains to the current TB epidemic in Tibet. Although we found a very high proportion of clustering and inferred transmission of drug-resistant strains, it is likely that these are underestimates, because our sample contained only about one-fifth of the culture-confirmed TB patients in Tibet during only the two short periods of time included in the study.

In conclusion, the high proportion of clustering suggests that there is considerable transmission of drug resistant *M. tuberculosis* in Tibet. The expansion of INH resistant clades from the 1970’s through the 1990’s and their increased propensity to acquire RIF resistance likely contributed to the current MDR-TB burden. Pragmatic strategies should be implemented for the early detection and effective treatment of both INH and RIF resistant strains.

## Contribution

QJ, KLW, and QG designed the study. HCL, LS, MG, XQZ, and JW collected the samples and data. QJ, QYL, and JEP analysed the data. TGC guided the genomic analysis. JRG and CGY suggested in the epidemiological analysis. QJ, HET, QG drafted the article. All the authors contributed to the manuscript and reviewed the final version.

## Data Availability

Sequencing reads are deposited in the NCBI (EMBL-EBI) under study accession number SRP276868.

## Acknowledgements

We thank the physicians and technicians of Tibetan CDC and Chinese CDC for the collection, culture and storage of clinical samples, and the conduction of the epidemiological review.

## Funding

This work was supported by the National Major Science and Technology Project of China [2017ZX10201302-006 and 2018ZX10715012-005 to QG, and 2018ZX10101002 to KLW]; and the Natural Science Foundation of China [91631301 and 81661128043 to QG].

## Disclaimer

The funders had no role in study design, data collection and analysis, decision to publish, or preparation of the manuscript.

## Competing interests

We declare no competing interests.

## Patient consent for publication

Not required.

## Ethics approval

Ethical approval was obtained from the Ethical Review Board of the Chinese Center for Disease Control and Prevention.

## Data availability statement

Sequencing reads are deposited in the NCBI (EMBL-EBI) under study accession number SRP276868.

## Notes

### Competing Interest Statement

The authors have declared no competing interest.

