## Supplemental for "Evolutionary trajectories and transmission dynamics of multidrug-resistant *Mycobacterium tuberculosis* in Tibet, China"

**Supplementary materials for the manuscript "Evolutionary trajectories and transmission dynamics of multidrug-resistant *Mycobacterium tuberculosis* in Tibet, China" by Q. Jiang et al.**

**Suppl. Table 1. Demographic and clinical characteristics of sampled tuberculosis (TB) patients, stratified by strain lineages**

| | Total N=576 (100%) | Lineage 3 N=16 (2.8%) | Lineage 4 N=36 (6.3%) | Lineage 2 N=524 (91.0%) | $\chi^2$ | P value |
|---|---|---|---|---|---|---|
| Location, n (%) | | | | | 1.23 | 0.539 |
| Lhasa | 300 (52.1) | 7 (43.8) | 16 (44.4) | 277 (52.9) | | |
| Other | 261 (45.3) | 8 (50.0) | 19 (52.8) | 234 (44.7) | | |
| Sample year | | | | | 5.93 | 0.204 |
| 2006 | 177 (30.7) | 2 (12.5) | 11 (30.6) | 164 (31.3) | | |
| 2009 | 172 (29.9) | 9 (56.3) | 11 (30.6) | 152 (29.0) | | |
| 2010 | 227 (39.4) | 5 (31.3) | 14 (38.9) | 208 (39.7) | | |
| TB history | | | | | 0.26 | 0.878 |
| New | 273 (47.4) | 8 (50.0) | 18 (50.0) | 247 (47.1) | | |
| Retreated | 288 (50.0) | 7 (43.8) | 17 (47.2) | 264 (50.4) | | |
| Genetic resistance, n (%) | | | | | 16.42 | 0.173 |
| Pan-susc. | 228 (39.6) | 13 (81.3) | 15 (41.7) | 200 (38.2) | | |
| Hr-TB | 48 (8.3) | 2 (12.5) | 2 (5.6) | 44 (8.4) | | |
| Rr-TB | 44 (7.6) | 0 (0) | 3 (8.3) | 41 (7.8) | | |
| Other resistance | 20 (3.5) | 0 (0) | 1 (2.8) | 19 (3.6) | | |
| MDR-TB | 187 (32.5) | 1 (6.3) | 10 (27.8) | 176 (33.6) | | |
| pre-XDR | 41 (7.1) | 0 (0) | 4 (11.1) | 37 (7.1) | | |
| XDR-TB | 8 (1.4) | 0 (0) | 1 (2.8) | 7 (1.3) | | |
| Genetic clustering, n (%) | | | | | | |
| 1-SNP | 105 (18.2) | 0 (0) | 3 (8.3) | 102 (19.5) | 6.47 | 0.039 |
| 5-SNP | 216 (37.5) | 2 (12.5) | 8 (22.2) | 206 (39.3) | 8.59 | 0.014 |
| 12-SNP | 311 (54.0) | 2 (12.5) | 12 (33.3) | 297 (56.7) | 18.80 | <0.001 |
| 20-SNP | 409 (71.0) | 13 (81.3) | 12 (33.3) | 384 (73.3) | 26.95 | <0.001 |
| 50-SNP | 509 (88.4) | 13 (81.3) | 17 (47.2) | 479 (91.4) | 64.81 | <0.001 |

Abbreviations: IQR=interquartile range; Pan-susc. = pan-susceptible; Hr-TB = isoniazid-resistant rifampicin-susceptible tuberculosis; Rr-TB = isoniazid-susceptible rifampicin-resistant tuberculosis; Other-resi. = susceptible to isoniazid and rifampicin, resistant to other anti-tuberculosis drugs; MDR = multidrug resistant (resistant to both isoniazid and rifampicin); Pre-XDR = pre-extremely drug resistant; XDR = extremely drug resistant.

**Suppl. Table 2. Number of strains with acquired and transmitted resistance to anti-tuberculosis drugs**

| Drug | # resistant strains | # strains of acquired resistance | # strains of transmitted resistance | % strains with transmitted mutation | # arisen events | # unique branches | # clusters | % events with transmission | Mean transmission size | Largest transmission size |
|---|---|---|---|---|---|---|---|---|---|---|
| SM | 231 | 40 | 191 | 82.7% | 70 | 40 | 30 | 42.9% | 6.4 | 41 |
| INH | 284 | 63 | 221 | 77.8% | 110 | 75 | 35 | 31.8% | 6.3 | 58 |
| RIF | 280 | 106 | 174 | 62.1% | 161 | 105 | 56 | 34.8% | 3.1 | 10 |
| EMB | 170 | 65 | 97 | 57.1% | 136 | 104 | 32 | 23.5% | 3.0 | 6 |
| PZA | 121 | 58 | 63 | 52.1% | 76 | 58 | 18 | 23.7% | 3.5 | 7 |
| FQ | 55 | 31 | 24 | 43.6% | 39 | 31 | 8 | 20.5% | 3.0 | 4 |
| AMK | 8 | 5 | 3 | 37.5% | 6 | 5 | 1 | 16.7% | 3.0 | 3 |
| CPM | 8 | 5 | 3 | 37.5% | 6 | 5 | 1 | 16.7% | 3.0 | 3 |
| KAN | 10 | 5 | 5 | 50.0% | 7 | 5 | 2 | 28.6% | 2.5 | 5 |
| ETO | 53 | 17 | 36 | 67.9% | 24 | 17 | 7 | 29.2% | 5.1 | 12 |
| PAS | 75 | 3 | 72 | 96.0% | 9 | 3 | 6 | 66.7% | 12.0 | 58 |
| LZD | 0 | - | - | - | - | - | - | - | - | - |
| BDQ | 2 | 2 | 0 | 0.0% | 2 | 2 | 0 | 0.0% | - | - |
| CLF | 2 | 2 | 0 | 0.0% | 2 | 2 | 0 | 0.0% | - | - |

Abbreviations: SM, streptomycin; INH, isoniazid; RIF, rifampicin; EMB, ethambutol; PZA, pyrazinamide; FQ, fluroquinolones; AMK, amikacin; CPM, capreomycin; KAN, kanamycin; ETO, ethionamide; PAS, para-aminosalicylic acid; LZD, linezolid; BDQ, bedaquiline; CLF, clofazimine.

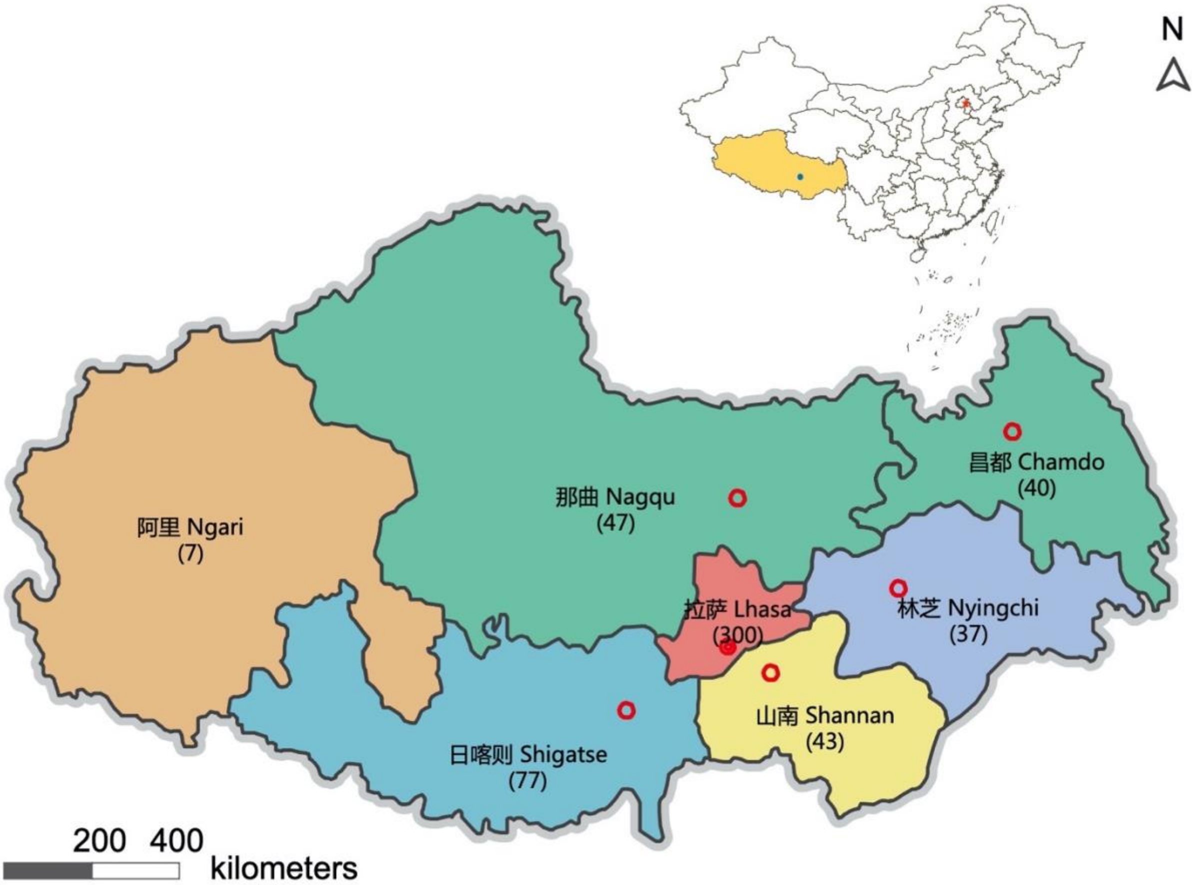


**Suppl. Figure 1. Districts of Tibet that were sources of the strains sampled in this study.** The number of samples collected from each district is shown in the brackets following the district name.

### Clade A

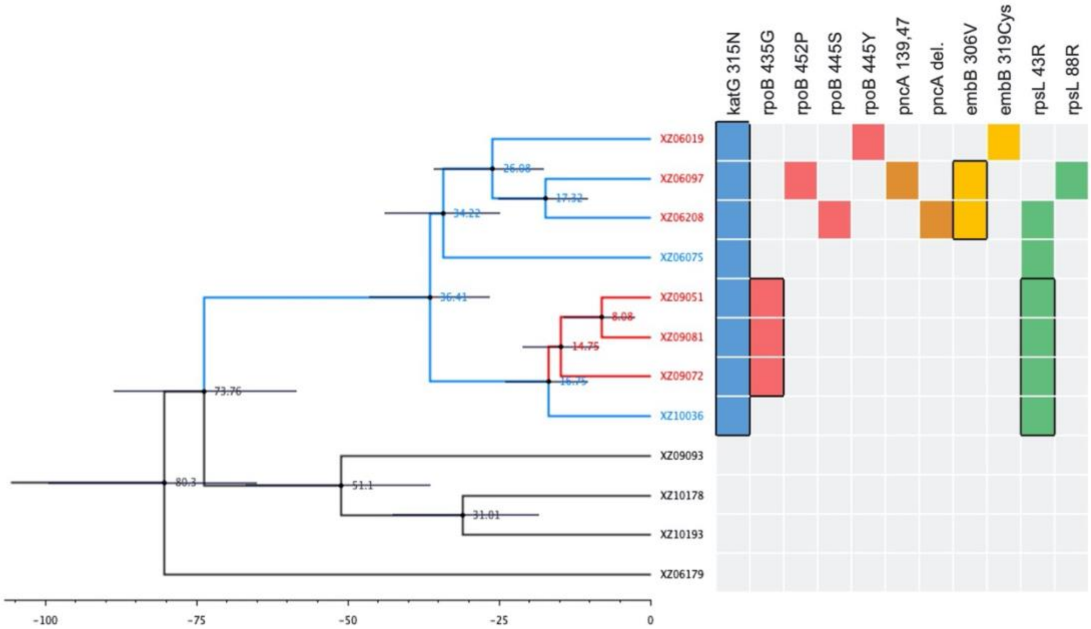


### Clade B

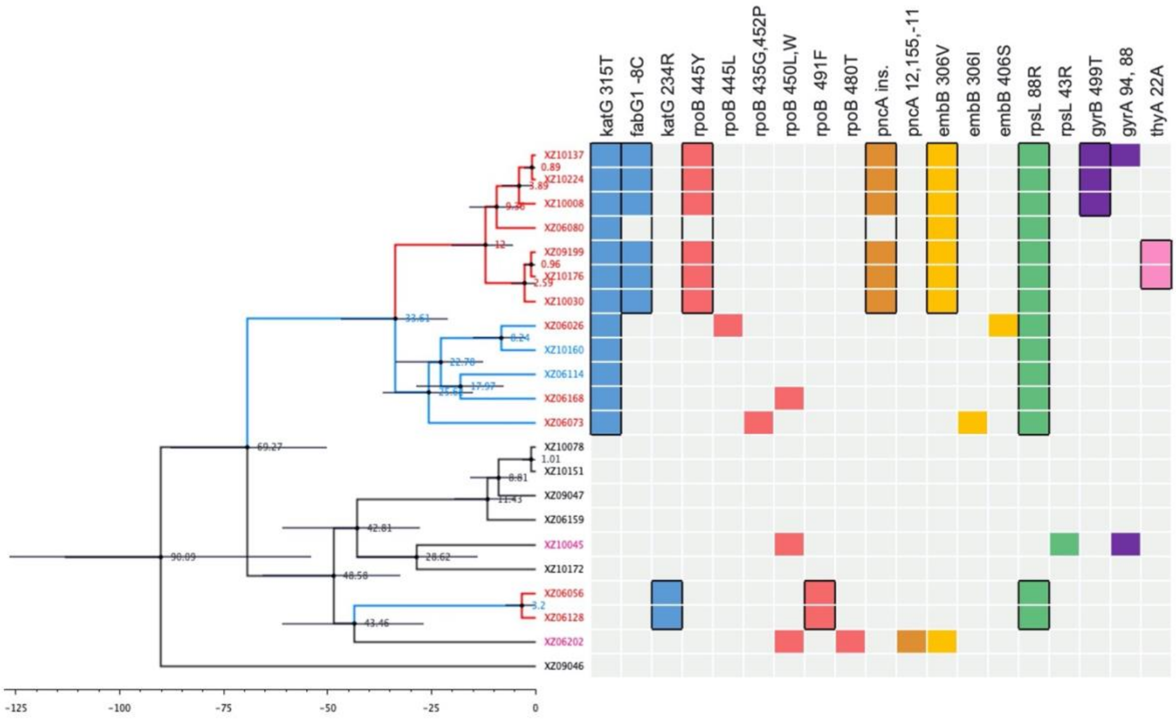

### Clade D

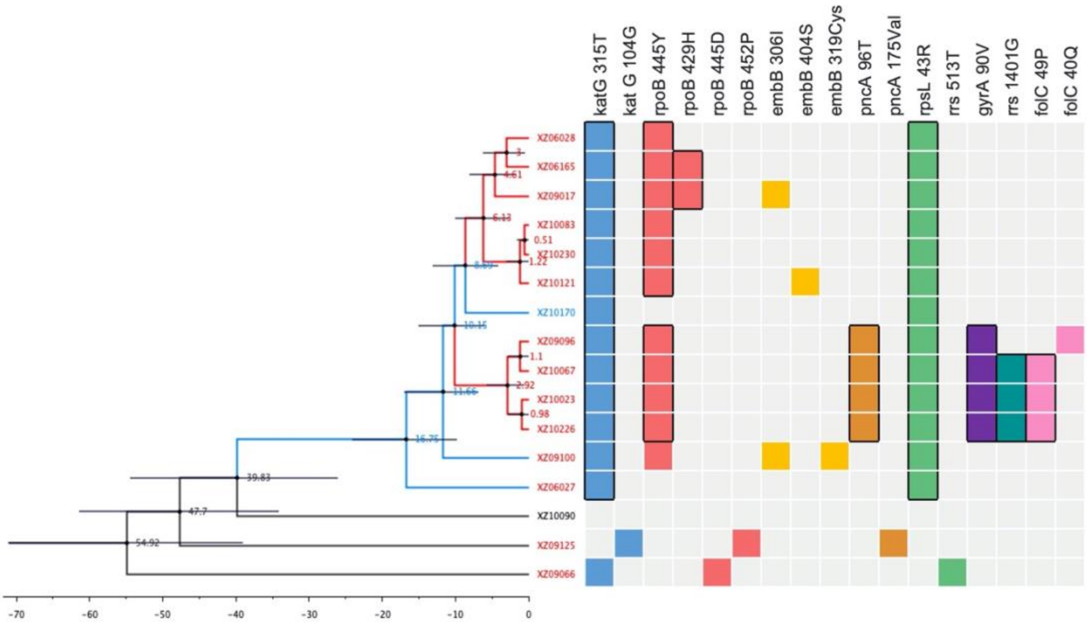


### Clade E

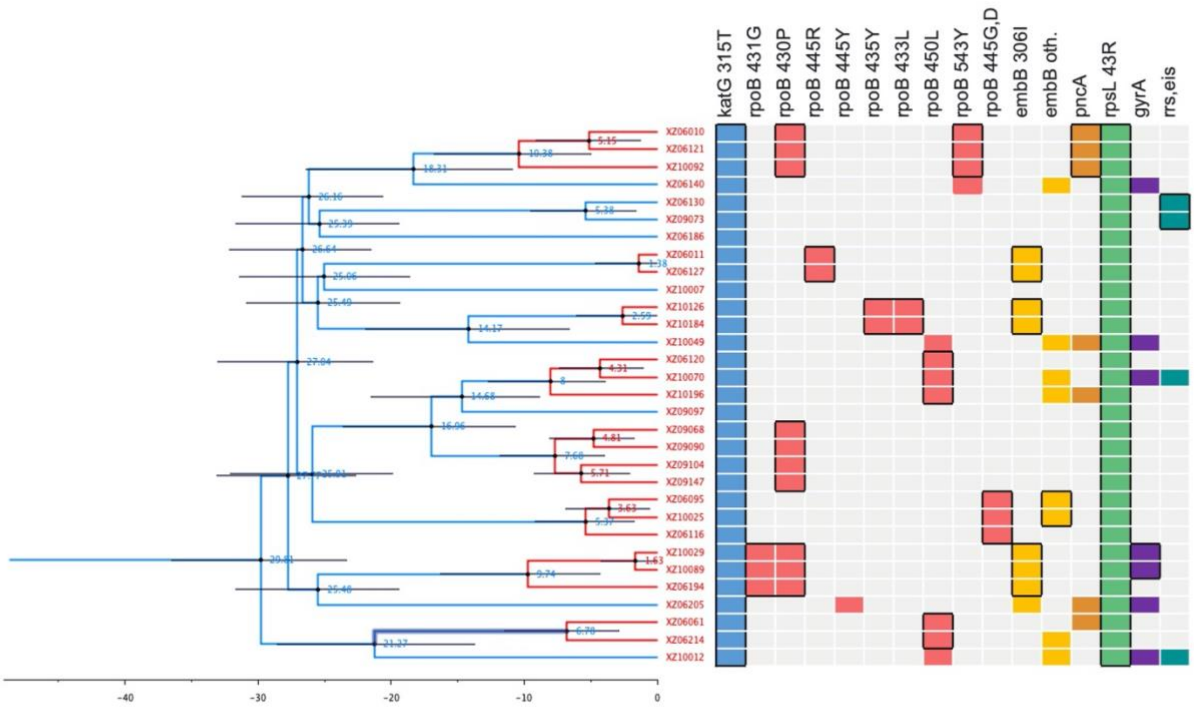

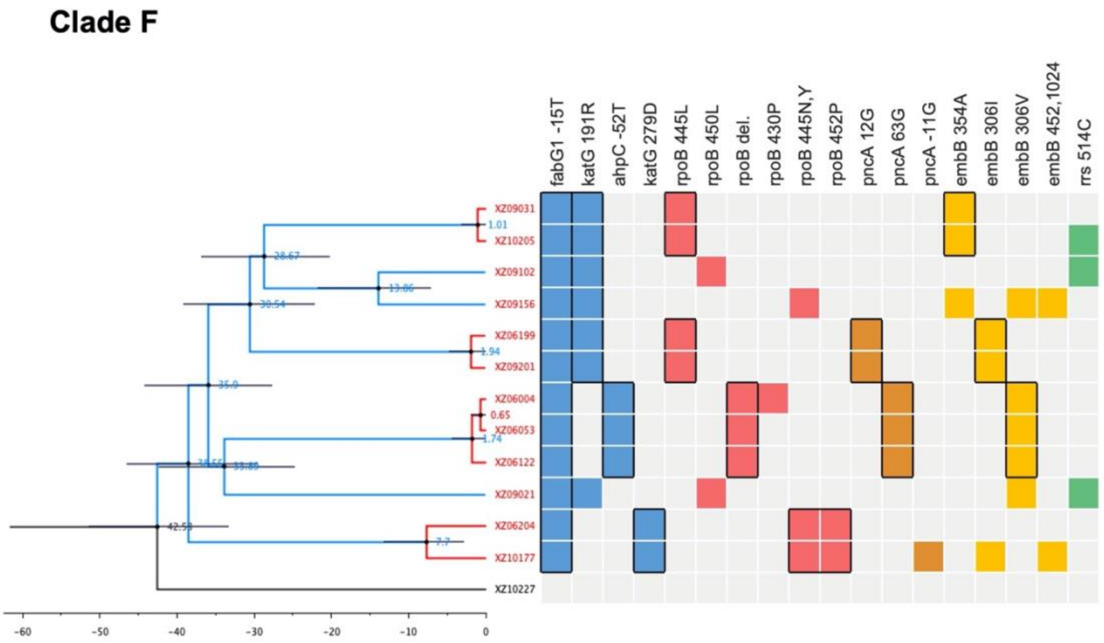


**Suppl. Figure 2. Trajectories of resistance evolution in major clades.** In each subfigure, on the left is the dated phylogeny tree of the clade, with blue and red branches showing transmitted resistance to isoniazid (INH) and rifampicin (RIF) respectively. Blue or red labels indicate INH-resistant or MDR strains. The right panels for each clade show the presence of drug resistance mutations, with different colours indicating resistance to the corresponding drugs.
